# Evidence for *GRN* as part of a neuroinflammatory mechanism connecting common neurodegenerative diseases: Genomic data analysis within an open science framework provides evidence connecting *GRN* to generalizable neurodegeneration

**DOI:** 10.1101/2020.12.04.20243600

**Authors:** Mike A. Nalls, Cornelis Blauwendraat, Lana Sargent, Dan Vitale, Hampton Leonard, Hirotaka Iwaki, Yeajin Song, Sara Bandres-Ciga, Kevin Menden, Faraz Faghri, Peter Heutink, Mark R. Cookson, Andrew B. Singleton

**Affiliations:** Laboratory of Neurogenetics, National Institute on Aging, National Institutes of Health, Bethesda, MD, USA 20892; Center for Alzheimer’s and Related Dementias, National Institutes of Health, Bethesda, MD, USA 20892; Data Tecnica International LLC, Glen Echo, MD, USA 20812; German Center for Neurodegenerative Diseases (DZNE), Tuebingen, Germany

**Author notes:** Corresponding author: Mike A. Nalls, PhD [ ].

## Abstract

**Background:** Previous research using genome wide association studies (GWAS) has identified variants that may contribute to lifetime risk of multiple neurodegenerative diseases. However, whether there are common mechanisms that link neurodegenerative diseases is uncertain. Here, we focus on one gene, *GRN*, encoding progranulin, and the potential mechanistic interplay between genetic risk, gene expression in the brain and inflammation across multiple common neurodegenerative diseases.

**Methods:** We utilized GWAS, expression quantitative trait locus (eQTL) mapping and Bayesian colocalization analyses to evaluate potential causal and mechanistic inferences. We integrate various molecular data types from public resources to infer disease connectivity and shared mechanisms using a data driven process.

**Findings:** eQTL analyses combined with GWAS identified significant functional associations between increasing genetic risk in the *GRN* region and decreased expression of the gene in Parkinson’s, Alzheimer’s and amyotrophic lateral sclerosis. Additionally, colocalization analyses show a connection between blood based inflammatory biomarkers relating to platelets and *GRN* expression in the frontal cortex.

**Interpretation:** *GRN* expression mediates neuroinflammation function related to general neurodegeneration. This analysis suggests shared mechanisms for Parkinson’s, Alzheimer’s and amyotrophic lateral sclerosis.

**Funding:** National Institute on Aging, National Institute of Neurological Disorders and Stroke, and the Michael J. Fox Foundation.

## INTRODUCTION

Alzheimer’s disease (AD), Parkinson’s disease (PD) and amyotrophic lateral sclerosis (ALS) are thought to be three distinct disorders representative of the common neurodegenerative disease (NDD) spectrum, with other less common conditions manifesting as combinatorial versions of these symptomatologies. While common lifestyle and environmental factors have been suggested to contribute to all three disorders, well-powered genome wide association studies (GWAS) generally indicate a distinct rather than shared set of chromosomal loci that contribute to lifetime disease risk.

Mutations in the *GRN* gene lead to decreased expression of the encoded protein, progranulin, and an increased risk of frontotemporal dementia (FTD) ^1–5^. The *GRN* locus has been nominated as contributing to the lifetime risk of several NDDs to varying extents ^6–9^. This locus is therefore a potential exception to the general rubric that NDDs have distinct genetic architecture.

Prior attempts to confirm single gene risks across NDDs have been limited to simple association studies and meta-analyses in small sample series ^10^. Here we apply post-GWAS methods to evaluate whether the *GRN* locus reliably contributes to risk of AD, PD and ALS in order to infer potential shared mechanistic consequences.

## METHODS

GWAS summary statistics were extracted from the three largest studies of AD, PD and ALS in the public domain for the chromosomal region surrounding *GRN*, defined as 500 kilobases from the 5’ and 3’ gene boundaries ^6–8^. To test the specific hypothesis that decreased expression of *GRN* is associated with increased risk of NDDs, we located a large dataset for brain gene expression that includes a meta-analysis of brain derived gene expression across multiple cohorts and brain regions, statistically accounting for sample overlap across studies ^11^.

To generate functional inferences relating to the role of *GRN* in gene expression for these three archetypes of NDD, we used summary-data-based Mendelian randomization (SMR) ^12^. SMR was carried out using default settings with the linkage disequilibrium (LD) reference dataset of over 30,000 samples described previously ^6^. Initial analyses were limited to testing the association with *GRN* and the three diseases, and *post-hoc* analyses were then applied to ascertain any other possible associations within the one megabase region surrounding the gene of interest. Associations were considered valid if P from the multi-SNP (single nucleotide polymorphism) test in SMR was less than 0.01 after multiple test correction. This threshold denotes a significant association between local SNPs and changes in expression that are functionally connected to disease etiology. When expanding past the *GRN* gene itself, we excluded SMR associations if the P for the heterogeneity in dependent instruments (HEIDI) test was less than 0.01, which would suggest potential violations of the inherent assumptions of the causal inference model.

To fine-map signals, we ran the approximate Bayes factor fine-mapping routine within the R package ‘coloc’ for each trait separately using all P values within the region of interest as an input ^13–15^. Null results for localizing a putative functional variant led us to not attempt any cross trait colocalization for these diseases. However, to gain further insight, we did mine aggregate colocalization data for *GRN* from the OpenTargets database (accessed November 23rd, 2020) ^16^.

## DATA AND CODE

Available here.

## RESULTS

We first examined associations between genomic variation at *GRN* and all three diseases of interest. The strongest effect was with PD, where a single standard deviation increase above the population mean genetic risk for PD at this locus was associated with a 0.1650 standard deviation decrease in expression of *GRN* in the brain (SE = 0.0354, P = 1.67E-07). A similar direction and magnitude of effect is seen in ALS (beta = −0.1038, SE 0.0411, P = 8.36E-03), and a smaller but significant effect seen in AD (beta = −0.0230, SE = 0.0064, P = 1.53E-03). The results are detailed in Table 1 and Figure 1 below.

**Table 1:** Summary of local SMR analyses using brain expression data and GWAS estimates for AD, ALS and PD.

| Symbol | Gene ID | Disease | N SNPs | Beta | SE | Multi-SNP P | HEIDI P |
| --- | --- | --- | --- | --- | --- | --- | --- |
| *GRN* | ENSG00000030582 | PD | 14 | -0.1650 | 0.0354 | 1.67E-07 | 0.0471 |
| *GJC1* | ENSG00000182963 | PD | 20 | -0.1925 | 0.0512 | 1.09E-04 | 0.3489 |
| *ASB16-AS1* | ENSG00000267080 | PD | 20 | 0.0805 | 0.0237 | 5.57E-04 | 0.0146 |
| *HIGD1B* | ENSG00000131097 | PD | 20 | 0.1890 | 0.0579 | 1.10E-03 | 0.1037 |
| *GRN* | ENSG00000030582 | AD | 18 | -0.0230 | 0.0064 | 1.53E-03 | 0.7770 |
| *GRN* | ENSG00000030582 | ALS | 18 | -0.1038 | 0.0411 | 8.36E-03 | 0.9823 |

**Figure 1:**
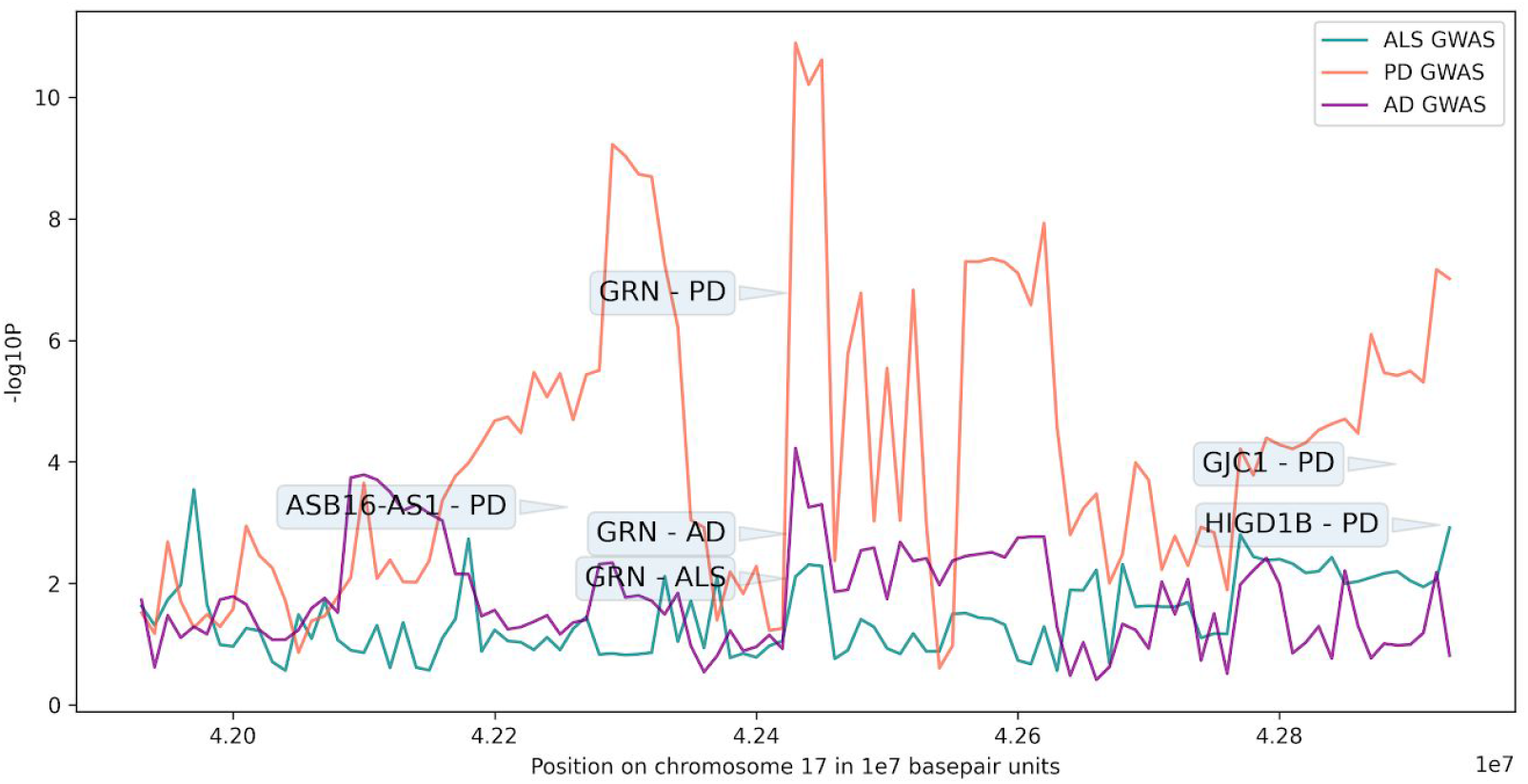
Graphical summary of SMR and GWAS analysis significance in the region per disease. To better display GWAS summary statistics, we only represent the most significant value per 10 kilobase window to avoid overplotting and improve clarity. The annotated squares represent the SMR results for genes of interest per disease. The y-axis represents negative log10 P values for each association, the x-axis represents positions on chromosome 17 (in 1E07 base pair units).

Fine-mapping and colocalization across these diseases at *GRN* failed to identify any single variant with posterior probabilities of causality greater than 0.80. Posterior probabilities for the null hypothesis, which relates to the likelihood of no quantifiable causal variant(s) neared this threshold, were 4.92E-06 in PD, 0.8743 in AD and 0.9142 in ALS. The top nominated SNP for PD (rs850738 in *FAM171A2*) only had a posterior probability of 0.1946 for being the causal variant, which is considered extremely weak evidence for causality. Therefore, no conclusive fine-mapping conclusion can be made from our GWAS derived data. We also failed to support a causal role for the nearby gene *FAM171A2* based on eQTL data.

Mining the OpenTargets database shows 55 associations suggesting colocalization evidence between GWAS studies and gene expression estimates at *GRN*. Out of these 55 putative causal associations, the overwhelming majority (80%) were related to platelet derived GWAS, with an additional 4 of the 55 associations related to white blood cell GWAS. This is a major overrepresentation of blood and potential inflammatory traits compared to others at this locus. These inflammatory related traits from blood (platelet distribution width, platelet count, mean platelet volume and thrombocyte volume) are all colocalized with variants related to changes in brain cortex expression ^17–19^.

## DISCUSSION

Here, we report strong causal functional inferences that connect genetic risk of three common NDDs to gene expression changes in the brain for *GRN*. This is the first study to link *GRN* to multiple NDDs and to suggest a gene-level mechanistic connection across a spectrum of NDDs. We additionally used a public resource, OpenTargets, to provide further mechanistic insights suggesting that the risk at this locus may be related to the body’s immune response to injury through inflammation and potentially further brain-related etiological effects.

The association between *GRN* and FTD is well documented in the literature. Granulin encodes the progranulin protein (PGRN) which can be cleaved into 7.5 granulin domains. Whereas PGRN has anti-inflammatory properties, granulins display pro-inflammatory properties (reviewed in DOI 10.1007/s12035-012-8380-8). A clear connection between decreased *GRN* expression and increased neuronal inflammation processes such as neutrophil migration and response to interleukins was seen in post mortem brain tissue of *GRN* mediated FTD and not in *MAPT* mediated FTD ^20^. Upregulated processes include NF-kappa-B (NFkB) signaling, as well as genes involved in tumour necrosis factor (TNF) production. In addition, extracellular matrix (ECM) pathways and matrix metalloproteases (MMPs) are up-regulated. Mediation of the inflammatory response involves proteolytic processing of anti-inflammatory GRN into pro-inflammatory granulins by the serine proteases neutrophil elastase, proteinase 3 and some MMPs (reviewed in DOI 10.1007/s12035-012-8380-8). In mouse models of ALS inhibition of the MMPs, MMP2 and MMP9 could indeed prolong survival and reduce symptoms. This prior data provides evidence for the potential role of inflammatory mediated responses driven by *GRN* with shared etiology for multiple NDDs. Identification of common pathogenic mechanisms across the different NDDs couldhelp explain shared clinical symptoms and establish more accurate genotype-phenotype correlations.

There are limitations to the current analyses that derive largely from data availability. As we used GWAS summary statistics, we did not have sufficient deep sequencing data for *GRN* in these diseases to accurately fine-map the locus. Additionally, due to sample size constraints, we were not able to include data from FTD subtypes or Lewy body dementia in our analyses. Finally, we do not have sufficient data to quantify the associations seen in diverse genetic ancestries but hope to evaluate this in the future. Finally, well powered GWAS in pathology confirmed samples would benefit future research and allow us to exclude to a degree potential artifacts related to misdiagnosis of NDDs.

## Data Availability

Available here [https://drive.google.com/file/d/1VVYpnhK00-R4Ah9MROcusF1U0hgjdIfx/view?usp=sharing].

https://drive.google.com/file/d/1VVYpnhK00-R4Ah9MROcusF1U0hgjdIfx/view?usp=sharing

## CONTRIBUTORS

MAN, CB, LS, DV, HL, HI,YS, SBC, KM, FF, PH, MC and AS contributed to the concept and design of the study, were involved in the acquisition of data, data generation, and data cleaning, the analysis and interpretation of data, and contributed to the drafting of the article and revising it critically.

## ACKNOWLEDGEMENTS

This work was supported in part by the Intramural Research Program of the National Institute on Aging and National Institute of Neurological Disorders and Stroke (project number Z01-AG000949-02).

## REFERENCES

1. Rademakers, R. et al. Phenotypic variability associated with progranulin haploinsufficiency in patients with the common 1477C→T (Arg493X) mutation: an international initiative. The Lancet Neurology vol. 6 857–868 (2007).

2. Goedert, M. & Spillantini, M. G. Frontotemporal lobar degeneration through loss of progranulin function. Brain vol. 129 2808–2810 (2006).

3. Cruts, M. & Van Broeckhoven, C. Loss of progranulin function in frontotemporal lobar degeneration. Trends in Genetics vol. 24 186–194 (2008).

4. Eriksen, J. L. & Mackenzie, I. R. A. Progranulin: normal function and role in neurodegeneration. J. Neurochem. 104, 287–297 (2008).

5. Le Ber, I. et al. Progranulin null mutations in both sporadic and familial frontotemporal dementia. Hum. Mutat. 28, 846–855 (2007).

6. Nalls, M. A. et al. Identification of novel risk loci, causal insights, and heritable risk for Parkinson’s disease: a meta-analysis of genome-wide association studies. Lancet Neurol. 18, 1091–1102 (2019).

7. Jansen, I. E. et al. Genome-wide meta-analysis identifies new loci and functional pathways influencing Alzheimer’s disease risk. Nat. Genet. 51, 404–413 (2019).

8. Nicolas, A. et al. Genome-wide Analyses Identify KIF5A as a Novel ALS Gene. Neuron 97, 1268–1283.e6 (2018).

9. Bellenguez, C. et al. Large meta-analysis of genome-wide association studies expands knowledge of the genetic etiology of Alzheimer’s disease and highlights potential translational opportunities. medRxiv 2020.10.01.20200659 (2020).

10. Noyce, A. J. et al. The Parkinson’s Disease Mendelian Randomization Research Portal. Mov. Disord. 34, 1864–1872 (2019).

11. Qi, T. et al. Identifying gene targets for brain-related traits using transcriptomic and methylomic data from blood. Nat. Commun. 9, 2282 (2018).

12. Zhu, Z. et al. Integration of summary data from GWAS and eQTL studies predicts complex trait gene targets. Nat. Genet. 48, 481–487 (2016).

13. Plagnol, V., Smyth, D. J., Todd, J. A. & Clayton, D. G. Statistical independence of the colocalized association signals for type 1 diabetes and RPS26 gene expression on chromosome 12q13. Biostatistics 10, 327–334 (2009).

14. Wallace, C. Statistical testing of shared genetic control for potentially related traits. Genet. Epidemiol. 37, 802–813 (2013).

15. Giambartolomei, C. et al. Bayesian test for colocalisation between pairs of genetic association studies using summary statistics. PLoS Genet. 10, e1004383 (2014).

16. Koscielny, G. et al. Open Targets: a platform for therapeutic target identification and validation. Nucleic Acids Res. 45, D985–D994 (2017).

17. UK Biobank — Neale lab. http://www.nealelab.is/uk-biobank.

18. Astle, W. J. et al. The Allelic Landscape of Human Blood Cell Trait Variation and Links to Common Complex Disease. Cell 167, 1415–1429.e19 (2016).

19. Consortium, T. G. & The GTEx Consortium. The GTEx Consortium atlas of genetic regulatory effects across human tissues. Science vol. 369 1318–1330 (2020).

20. Menden, K. et al. Integrated multi-omics analysis reveals common and distinct dysregulated pathways for genetic subtypes of Frontotemporal Dementia. Cold Spring Harbor Laboratory 2020.12.01.405894 (2020) doi:10.1101/2020.12.01.405894.

